# Prediction and stratification of longitudinal risk for chronic obstructive pulmonary disease across smoking behaviors

**DOI:** 10.1101/2023.04.04.23288086

**Authors:** Yixuan He, David C. Qian, James A. Diao, Michael H. Cho, Edwin K. Silverman, Alexander Gusev, Arjun K. Manrai, Alicia R. Martin, Chirag J. Patel

**Author notes:** Corresponding authors Correspondence to: Alicia R Martin, Analytic and Translational Genetics Unit, Massachusetts General Hospital, Richard B. Simches Research Center, 185 Cambridge Street, CPZN-6818, Boston, MA 02114, Chirag J Patel, 10 Shattuck St, Boston, MA 02215. Contributed equally (Alicia R. Martin, Chirag J. Patel).

## Abstract

Smoking is the leading risk factor for chronic obstructive pulmonary disease (COPD) worldwide, yet many people who never smoke develop COPD. We hypothesize that considering other socioeconomic and environmental factors can better predict and stratify the risk of COPD in both non-smokers and smokers. We performed longitudinal analysis of COPD in the UK Biobank to develop the Socioeconomic and Environmental Risk Score (SERS) which captures additive and cumulative environmental, behavioral, and socioeconomic exposure risks beyond tobacco smoking. We tested the ability of SERS to predict and stratify the risk of COPD in current, previous, and never smokers of European and non-European ancestries in comparison to a composite genome-wide polygenic risk score (PGS). We tested associations using Cox regression models and assessed the predictive performance of models using Harrell’s C index. SERS (C index = 0.770, 95% CI 0.756 to 0.784) was more predictive of COPD than smoking status (C index = 0.738, 95% CI 0.724 to 0.752), pack-years (C index = 0.742, 95% CI 0.727 to 0.756). Compared to the remaining population, individuals in the highest decile of the SERS had hazard ratios (HR) = 7.24 (95% CI 6.51 to 8.05, P < 0.0001) for incident COPD. Never smokers in the highest decile of exposure risk were more likely to develop COPD than previous and current smokers in the lowest decile with HR=4.95 (95% CI 1.56 to 15.69, P=6.65×10^−3^) and 2.92 (95%CI 1.51 to 5.61, P=1.38×10^−3^), respectively. In general, the prediction accuracy of SERS was lower in the non-European populations compared to the European evaluation set. In addition to genetic factors, socioeconomic and environmental factors beyond smoking can predict and stratify COPD risk for both non- and smoking individuals. Smoking status is often considered in screening; other non-smoking environmental and non-genetic variables should be evaluated prospectively for their clinical utility.

## INTRODUCTION

Chronic obstructive pulmonary disease (COPD), characterized by persistent obstruction to airflow in and out of the lungs, is the third leading cause of death globally^1^. While tobacco smoking is widely recognized as the single most important risk factor for COPD, it is now well-established that 20%-30% of COPD cases worldwide consist of never smokers^2,3^, and only 25% of continuous smokers will develop incident COPD^4,5^. This suggests that other risk factors such as non-smoking exposures and genetic markers also play important roles in pathogenesis.

Heritability estimates for COPD typically range between 20-50%^6,7^, and several large genome-wide association studies (GWAS) have uncovered significant genetic risk loci ^8–10^. Recently, a composite polygenic risk score (PGS) consisting of over 2 million genetic variants across the genome has been demonstrated to predict incident COPD and age of diagnosis better than previously published genetic risk scores^11–14^. However, a significant proportion of phenotypic and disease variance is still unexplained and likely attributable to environmental exposures^15^.

To date, non-genetic environmental studies of COPD have primarily focused on the relationships between a single or a small group of factors without considering the dense correlations of the exposome^16–18^. There does not exist any metric to summarize the cumulative effects of socioeconomic and environmental exposures beyond smoking for COPD. Previously, polyexposure risk scores which summarize the cumulative risk of many exposures have been constructed for other common diseases such as type 2 diabetes and cardiovascular disease and have provided more meaningful predictive performance and risk classification than single risk factors^19,20^. We hypothesize that a similar risk score accounting for socioeconomic and environmental factors beyond smoking will also improve COPD prediction and identify individuals with the highest risk of developing COPD.

COPD disproportionately affects individuals in ethnic minority groups – some of the strongest environmental risk factors for COPD, such as tobacco use and occupational exposures to fumes and chemicals, as well as heritability and susceptibility loci differ greatly in prevalence between populations^17,21–23^. Despite these differences, most studies have focused on individuals of European ancestry. In genetic studies where the reference population has consisted of individuals of European ancestry, the predictive performance of PGS is attenuated in non-European ancestry populations^24,25^. It is unclear whether this will also be true for environmental and socioeconomic factors.

In this study, we constructed and validated the COPD Socioeconomic and Environmental Risk Score (SERS) in a longitudinal cohort analysis that is conditional on smoking behaviors in the UK Biobank. We sought to determine whether SERS can predict and stratify disease risk across different smoking behaviors, especially among individuals who have never or rarely smoked. We evaluated our score in a held-out set consisting of multiple racial and ethnic groups to determine the generalizability of socioeconomic and environmental risk factors across populations.

## METHODS

### Study Cohort

The UK Biobank (UKB) is a large observational study of over half a million participants between 40-69 years of age at the time of recruitment between 2006 and 2010 ^26^. In our analysis, we excluded individuals who had a COPD diagnosis prior to the time of assessment, had missing diagnosis or followup time, were related, or had missing covariates. There were 320,115 individuals remaining. We used ancestry assignments from the Pan-UK Biobank (PanUKBB) project^27^, which was downloaded through the UKB portal as Return 2442. Based on the ancestry assignments, we identified 358,627 Europeans (EUR), 8,284 Central South Asians (CSA), 6,446 Africans (AFR), 2,641 East Asians (EAS), 1,578 Middle Easterners, and 970 Admixed Americans (AMR). In brief, PanUKBB conducted a pan-ancestry analysis of the UKB by comparing the genome of UKB participants against two large diverse global datasets, the 1000 Genome Project and the Human Genome Diversity Project, and assigned ancestry based on genetic similarity. The European participants were randomly divided into three subgroups, in a roughly 3:3:2: ratio (association testing N=113,714, derivation of SERS N=113,291, evaluation N=93,110). The evaluation subset, used for assessing the performances of the risk scores, also contained all non-European individuals. We used the entire association testing subgroup to conduct the initial exposure-wide association study on COPD. SERS was calculated for 84,998 individuals in the evaluation subgroup who had complete exposure responses for the final SERS factors.

### Phenotype Ascertainment

We classified COPD based on a combination of linked hospital admission records for International Classification of Disease (ICD) 9 codes of 490, 491, 492, 494, 496 ICD-10 codes of J41.X, J43.X, J44.X, J98.2, J98.3, having a forced expiratory volume (FEV1)/forced vital capacity (FVC) ratio of < 0.70, or having self-reported COPD in an interview. We used the earliest recorded time as the time of event in our analysis. In the full population, we identified 8,632 individuals diagnosed with COPD by self-report, 14,677 by ICD10 code, and 30 by ICD9 code. We excluded individuals who had a COPD diagnosis at the time of the first assessment. There were also 50,599 additional individuals who had an FEV1/FVC ratio of <0.70 at the time of assessment who were excluded from our study analysis.

### Socioeconomic and Environmental Risk Score Derivation and Validation

SERS captures the cumulative impact of socioeconomic, environmental and behavioral exposure risks. Individuals receive a score based on the weighted sum of many common non-genetic factors to which they may be exposed. Weights are determined by the strength of corresponding associations with the outcome of interest.

Methods for SERS derivation are previously described and are available through the R package PXStools^19,20^. In summary, we first conducted an exposure-wide association study (XWAS) for incident COPD in the derivation subgroup^28,29^. We then iterated through a LASSO-based stepwise selection procedure to identify independent features associated with longitudinal COPD development in the testing subgroup. We calculated the final SERS for the evaluation subgroup by taking the weighted sum of the exposure variables. In each step, we adjusted for sex, age, age^2^, age × sex, the first four principal components of genetic ancestry, smoking status, and pack-years.

The initial set of exposure variables we included were indicators of physiological state, environmental exposure, and self-reported behavior collected during the first assessment visit period (2006-2010). We wanted to construct a risk score separate of smoking effects, thus we did not consider any exposures in the “Smoking” category. Altogether, we started with 102 unique variables in total. Among these, we only considered variables that had less than 10% missingness, resulting in 83 variables for our pipeline. We processed our exposure data using the PHESANT software tool^30^. We excluded responses of “Prefer not to answer” and “Do not know”. For unordered categorical variables, the response with the largest number of participants was selected as the reference group. Individuals that responded “never smoked” were assigned a pack-year of zero. The final risk score contained 10 independent and significant exposure factors.

### Polygenic Score Derivation

We reconstructed a composite PGS for lung function and COPD consisting of roughly 2.5 million genetic variants^11^. In summary, Moll et al. derived the most comprehensive and accurate composite PGSs for COPD by training a logistic regression model based on two separate PGSs for forced expiratory volume in 1 second (FEV1) and FEV1/forced vital capacity (FVC) based on results from the genome-wide association studies of lung function from UKB and SpiroMeta. The composite PGS consisting of roughly 2.5 million SNPs identified individuals with elevated risk for moderate-to-severe COPD, emphysema subtypes associated with cigarette smoking, and radiographic patterns of reduced lung growth. The PGS has also been demonstrated to be associated with incident COPD and age of onset in large population-based cohorts^13,14^.

### Statistical Analysis

We conducted all analyses in R version 3.5.1. We placed individuals into bins by their PGS and SERS percentiles and calculated the prevalence of COPD within each bin as well as the hazard ratios for COPD in the top bins of PGS and SERS compared to the remaining individuals. We performed multivariable Cox proportional hazards regressions of COPD on combinations of risk scores and covariates. The base model contained only the covariates sex, age, age^2^, age × sex, and the first four principal components of genetic ancestry. The hazard ratios for membership in certain subgroups versus another subgroup were calculated by fitting a Cox regression model with a binary indicator variable. To measure the gross gene-environment correlation, we estimated the Pearson correlation coefficient between PGS and SERS. The standard errors of relative prediction accuracy were calculated by taking the absolute standard error multiplied by relative accuracy. To adjust for multiple tests, we used the “p.adjust” function of the base stats R package for Benjamini-Hochberg False Discovery Rate (FDR) adjustment.

## RESULTS

A schematic of our study design is shown in **Figure 1**. After excluding related individuals with missing information or who had previous/current COPD diagnoses, our study sample consisted of 320,115 individuals (median age 57, with 209,600 females [65.5%]).

**Figure 1:**
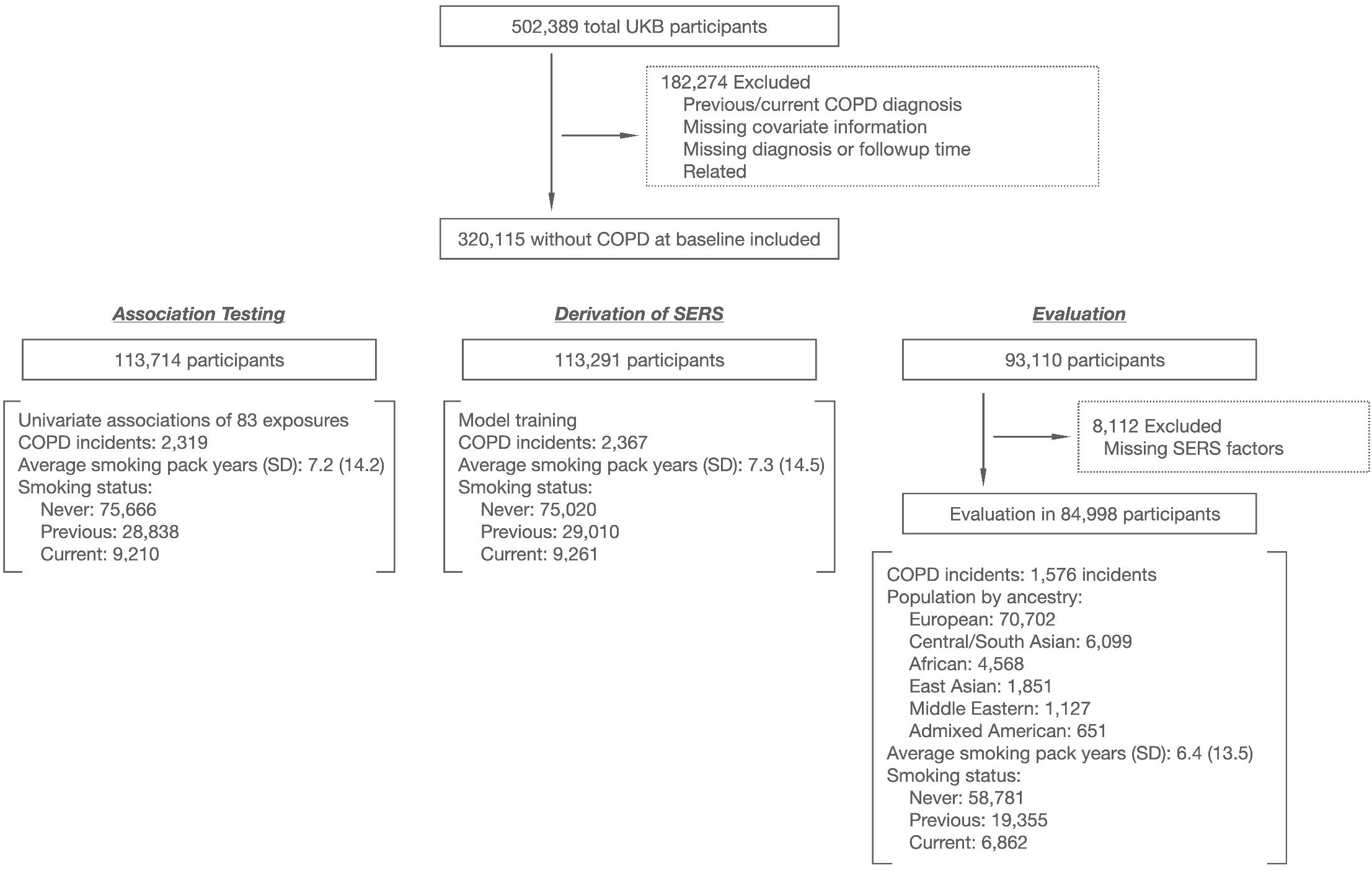
Study design. COPD: Chronic obstructive pulmonary disease. SERS: Socioeconomic and environmental risk score. UKB: UK Biobank. XWAS: eXposure wide association study.

We first tested univariate associations (XWAS)^29^ between 83 factors in the categories of categories “Sociodemographics”,’ Lifestyles and environment”, Residential air pollution”, and “Residential noise pollution” and COPD incidence. There were 26 factors that were significant (P<0.05) after correcting for multiple hypothesis testing (**Supplementary Figure 1**). We used the EWAS summary statistics to develop the COPD socioeconomic and environmental risk score (SERS). (**Supplementary Tables 1, Supplementary Figure 1**). After applying the PXStools algorithm, the final SERS for longitudinal COPD development consisted of 11 exposures: “Type of accommodation lived in”, “Own or rent accommodation lived in”, “Alcohol drinking status”, “Bread type”, “Current employment status”, “Nitrogen dioxide (2006)”, “Types of transport used”, “Types of physical activity in past four weeks”, “Major dietary changes in the past 5 years”, “Attendance/disability/mobility allowance”, “Time spent watching TV”. Since we were interested in developing a SERS that considered factors independent of smoking behaviors, we did not include smoking status or pack-years as an input exposure but instead adjusted for them in our association testing and SERS derivation. In the multivariable model, socioeconomic status and air pollution factors, such as having a disability allowance (HR=1.71, P < 0.0001), renting compared to owning (HR 1.66, P < 0.0001), and NO2 levels (HR=1.01, P=1.77×10^−4^), were most significantly associated with increased risk of COPD (**Supplementary Table 2**). Consuming white bread compared to multigrain (HR=1.14, P=8.10×10^−3^), being unemployed (1.49,p=0.0123), and being a previous alcohol drinker (HR=1.23, p=0.0224) were also significantly associated with increased risk of COPD. Walking compared to driving a car as the primary source of transportation was significantly associated with decreased risk of COPD (HR=0.790,p=7.22×10^−4^).

We first assessed COPD risk stratification by SERS and smoking behaviors (**Figure 2, Supplementary Figure 2**) in the European ancestry population (EUR). SERS, smoking status, and pack-years are all positively associated with COPD incidence in an additive manner (P < 0.0001)i. We binned individuals by SERS percentiles. Incidence of COPD spanned from 0.28%to 21.64% across SERS percentiles. Compared to the remaining population, individuals in the highest quintile and decile of SERS had hazard ratios (HR) of 5.2 5 (95% CI 4.73 to 5.84, P < 0.0001) and 7.24 (95% CI 6.51 to 8.05, P < 0.0001), respectively, for COPD. The HR of each SERS quintile compared to the first quintile in the EUR evaluation subset can be found in **Supplementary Tables 3**.

**Figure 2:**
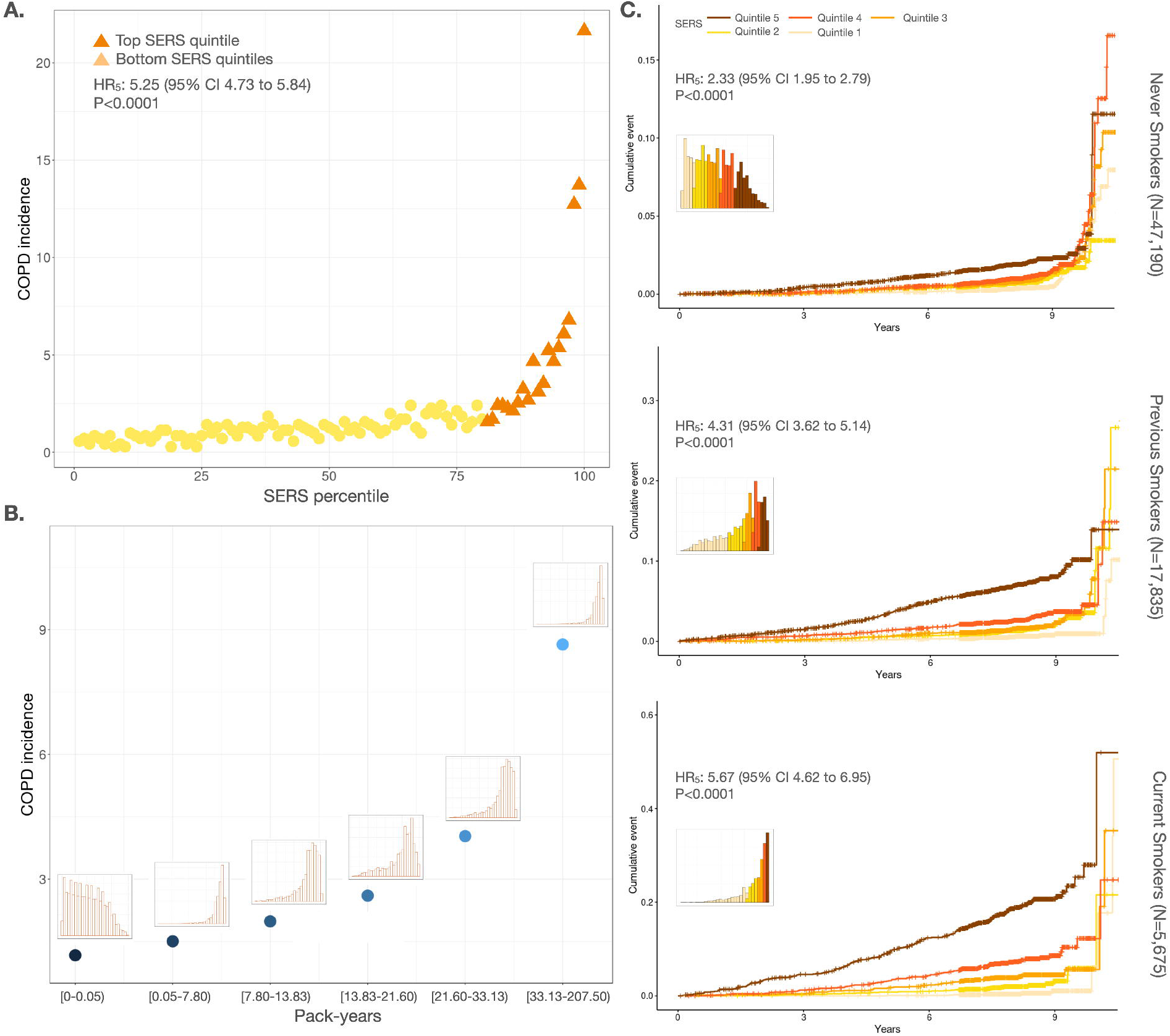
Disease stratification and prediction by SERS across smoking statuses. a) Incidence of COPD in each percentile of the evaluation set. The top quintile is colored in dark orange. b) COPD incidence for each pack-year quintile. The distribution of SERS for individuals in each quintile is shown above each point. c) Cumulative incidence plots for never smokers (top), previous smokers (middle), and current smokers (bottom) stratified by SERS (orange shades) quintiles. The distribution of SERS for each smoking status is shown in each panel.

Despite smoking being one of the strongest known predictors of COPD, not all individuals who smoke develop COPD, and many individuals who do not smoke develop COPD. In the EUR evaluation set, 548/47,190 (1.44 per 1000 person years) in never smokers, 504/17,835 (3.57 per 1000 person years) in previous smokers, and 373/5,675 (8.43 per 1000 person years) in current smokers had incident COPD. Current (HR=6.10, 95% CI 5.35 to 6.96, P<0.0001) and previous (HR=2.56, 95% CI 2.27 to 2.89, P<0.0001) smokers had a much higher risk of COPD compared to never smokers. Higher pack-years of smoking were also associated with a higher incidence of COPD and greater risk of COPD with an HR of 1.36 (95% CI 1.34 to 1.38, P<0.0001) per 10 pack-years smoked. SERS was modestly correlated with pack-years (r^2^=0.373, P<0.0001), but SERS was able to stratify COPD risk across smoking behaviors with better granularity. For each smoking status category (never smoker, past smoker, or current smoker), we binned individuals into percentiles by SERS. Regardless of smoking status, COPD incidence increased as SERS increased (**Supplementary Figure 3**). Between the highest and lowest SERS deciles, COPD incidence spanned 0.6-2.2% among never smokers, 0.6-9.5% among past smokers, and 0.5-21.0% among current smokers. We also estimated the 10 year cumulative incidence of COPD stratified by SERS (**Figure 2C**). In never smokers, previous smokers, and current smokers, individuals in the highest deciles of SERS had an HR of 2.40 (95% CI 1.94 to 2.99, P < 0.0001), 5.14 (95% CI 4.13 to 6.40, P < 0.0001) and 5.40 (95% CI 4.48 to 6.50, P < 0.0001), respectively, for developing COPD compared to the remaining population.

Having demonstrated the ability of SERS to stratify risk within smoking status categories, we then investigated if SERS is able to predict COPD risk across different smoking behaviors. Never smokers in the highest SERS decile had an HR of 4.95 (95% CI 1.56 to 15.69, P=6.65×10^−3^) and 2.92 (95%CI 1.51 to 5.61, P=1.38×10^−3^) compared to current smokers in the bottom decile and quintile of SERS, respectively. Never smokers in the highest SERS decile also had higher risks for COPD compared to previous smokers in the bottom decile (HR=4.54, 95% CI 2.39 to 8.60, P < 0.0001) and bottom quintile (HR=3.49, 95% CI 2.26 to 5.39, P < 0.0001) of SERS. We also found that in individuals who had future COPD incidence, one decile increase in SERS resulted in, on average, 0.26 years shorter time to disease (P=7.21×10^−4^) (**Supplementary Figure 4 and 5**).

We evaluated the performance of SERS to predict incident COPD and found a C index of 0.770 (95% CI 0.756 to 0.784) (**Figure 3**). The predictive ability of SERS was significantly higher than both smoking status (C index 0.738, 95% CI 0.725 to 0.752) and pack-years (C index 0.742, 95% CI 0.727 to 0.756). In the joint model (C index 0.771 95% CI 0.757 to 0.785), all three factors with significantly associated with COPD, with pack-years (P < 0.0001) being the most significant, followed by smoking status (P < 0.0001), being a current smoker (P < 0.0001), and being a previous smoker compared to never smoking (P=3.02×10^−2^). The performance of SERS was lower in predicting COPD in smoking status subgroups but higher than pack-years, with C indices for the SERS model of 0.656 (95% CI 0.630 to 0.681) in never smokers, 0.744 (95% CI 0.721 to 0.767) in previous smokers, and 0.777 (95% CI 0.756 to 0.798) in current smokers.

**Figure 3:**
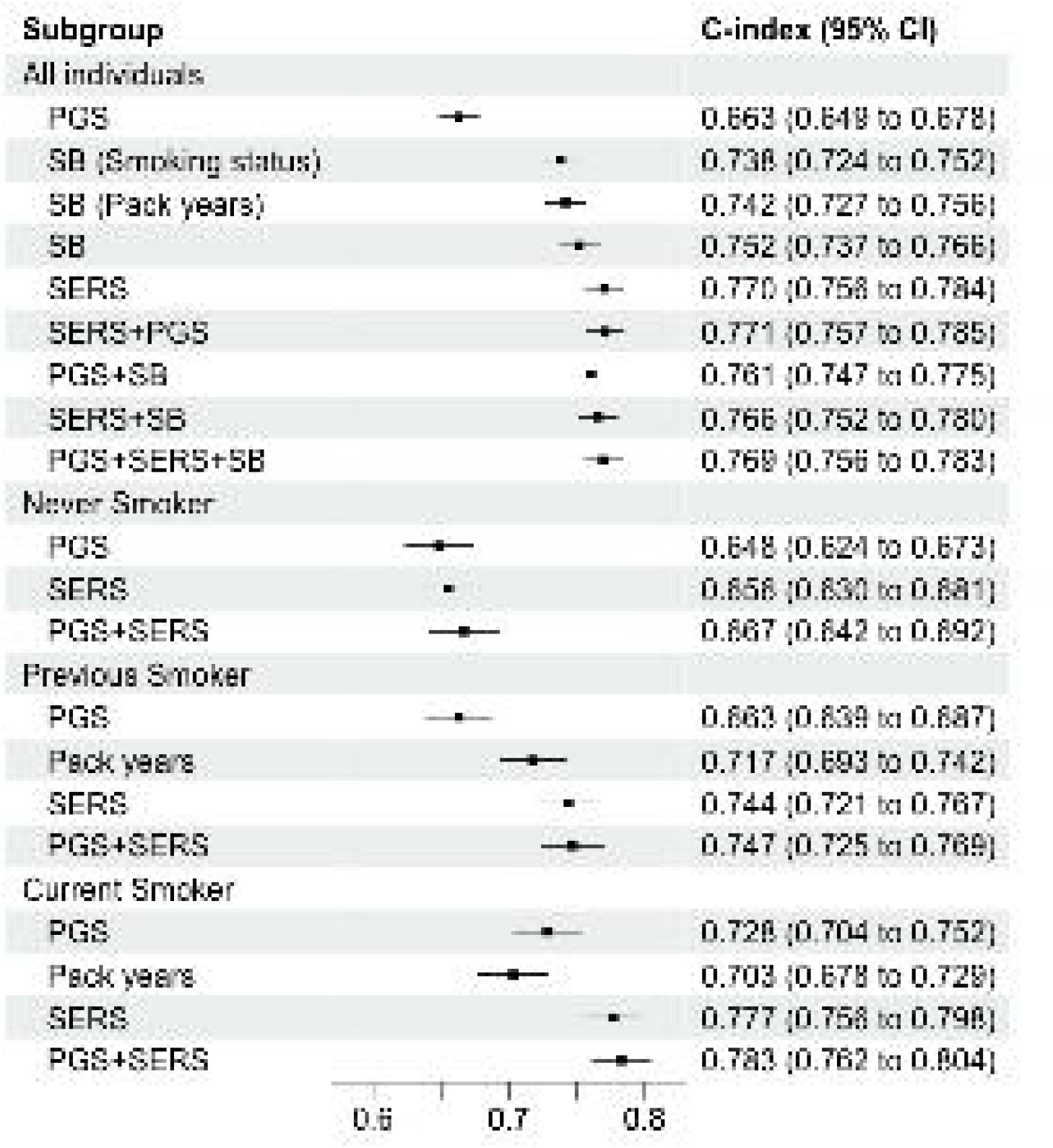
Performance of each prediction model. The C-indices and 95% confidence intervals for predicting COPD by various models across different smoking status subgroups. All models include baseline factors sex, age, age^2^, sex×age, and the first four principal components of genetic ancestry. SB: smoking behaviors, SERS: socioeconomic and environmental risk score, PGS: polygenic risk score.

To assess the complementarity and additivity of the SERS and polygenic risk, we computed a composite genome-wide polygenic risk score (PGS) from published weights of 2.5 million SNPS that is predictive of incident COPD and age of onset^11,13,14^. In our study population, we found that the PGS had lower predictive accuracy than smoking behaviors or SERS in the entire evaluation cohort (C index = 0.663, 95% CI 0.649 to 0.678) as well as within each smoking group (**Figure 2, Supplementary Figure 6**). The composite PGS is able to also stratify risk of COPD. Individuals in the top decile of PGS had an HR of 1.69 (95% CI 1.51 to 1.89, P<0.0001) compared to the rest of the population (**Supplementary Table 4**). In never smokers, previous smokers, and current smokers, individuals in the highest deciles of PGS had an HR of 1.74 (95% CI 1.38 to 2.18, P<0.0001), 1.66 (95% CI 1.25 to 2.21, P = 4.56×10^−4^) and 1.90 (95% CI 1.51 to 2.39, P<0.0001), respectively, for developing COPD compared to the remaining population. PGS was also unable to stratify non-smokers who had a higher risk of COPD than previous or current smokers.

To measure gross gene-environment correlation, we estimated the Pearson correlation coefficient between SERS and PGS. Socioeconomic and environmental factors were independent of genetic risks (P-value=0.3) in the EUR evaluation population. To further investigate genetic and environmental interactions in COPD, we classified individuals into five categories based on whether they were in the top or bottom quintiles of SERS and PGS: high SERS and high PGS, high SERS and low PGS, low SERS and high PGS, low SERS and low PGS, or none of the above. 209/2761 (7.6%) of individuals with both high SERS and PGS were later diagnosed with COPD, while 4/2755 (0.15%) of individuals with both low SERS and PGS were later diagnosed with COPD. Individuals with high SERS and high PGS had an HR of 4.80 (95% CI 4.14 to 5.56, P<0.0001) for COPD compared to the rest of the population (**Supplementary Figure 7**). To investigate how either SERS or PGS may identify risk not implicated by the other score, we compared individuals with high SERS and low PGS, and individuals with low SERS and high PGS. Individuals with high SERS and low PGS had an HR of 4.50 (95% CI 3.08 to 6.57, P<0.0001) for COPD compared to individuals with low SERS and high PGS, suggesting that low cumulative genetic risk may mediate high cumulative exposure risk. Of all individuals who were later diagnosed with COPD (1,380), there were 435 (31.5%) individuals with only high SERS,175 (12.7%) with high PGS, and 209 (15.1%) individuals with both high SERS and high PGS (**Supplementary Figure 8**).

Lastly, because COPD has known prevalence differences across populations^22,31^, we investigated differences in predictors of COPD between the European majority ancestry group versus the non-European ancestry minority groups. In the testing set, there were 14,296 total non-European ancestry individuals: 6,099 individuals of Central/South Asian (CSA) ancestry, 4,568 of African (AFR) ancestry, 1,851 of East Asian (EAS) ancestry, 1,127 of Middle Eastern (MID) ancestry, and 651 of Admixed American (AMR) ancestry. COPD incidence was much lower in these populations. There were 9 (1.78%) AMR incident cases, 38 (1.09 per 1000 person years) AFR cases, 61 (1.30 per 1000 person year) CSA cases, 19 (1.30 per 1000 person years) EAS cases, 20 (2.27 per 1000 person years) MID cases, and, for a total of 147 (1.34 per 1000 person year) incidents cases in non-European populations.

We calculated SERS for non-European ancestry populations using weights derived from the EUR reference population, which predicted COPD risk with a C index of 0.739 (95% CI 0.695 to 0.760) for SERS and PGS, respectively. We then randomly subsampled 1,500 individuals from each of the four largest ancestry groups: EUR, CSA, AFR, and EAS each for subsequent analyses. SERS had worse prediction in all three non-European population subgroups compared to the European ancestry subgroup. (**Figure 4a**). We investigated the distribution of smoking status (**Figure 4b**) and two SERS exposures, qualifications, and accommodations in the entire evaluation population (**Figure 4c-d**). The CSA population had the largest proportion (85.8%) of never smokers, followed by AFR (81.5%), EAS (84.1%), and EUR (66.7%). In the UKB population, CSA, AFR, and EAS ancestry populations consistently had the highest proportion of never alcohol drinkers and being in paid employment. The absolute values can be found in **Supplementary Table 5**.

**Figure 4:**
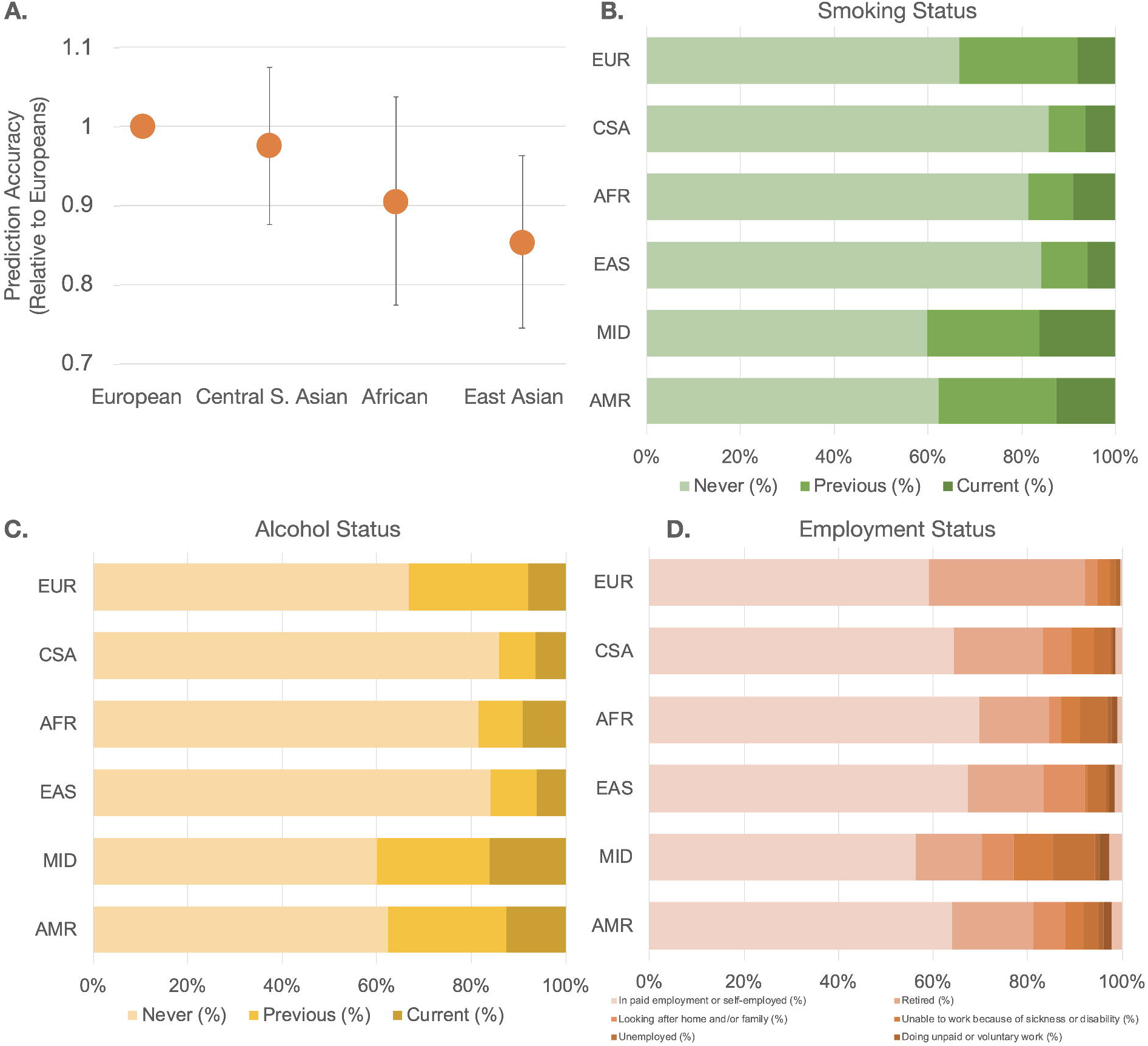
Prediction of COPD across ancestry groups by SERS. (a) Prediction accuracy of the top three non-European ancestry subgroups relative to European ancestry individuals with standard error bars. Distribution of (b) smoking status (left to right: Never, Previous, Current), © alcohol status (left to right: Never, Previous, Current), and (d) employment (left to right: In paid employment or self-employment, Retired, Looking after home and/or family, Unable to work because of sickness or disability, Unemployed, Doing unpaid or voluntary work, Full or part-time student, None of the above) across all ancestry groups.

## DISCUSSION

We developed a socioeconomic and environmental risk score (SERS) associated with time to COPD onset that is trained and evaluated on socioeconomic, environmental, and behavior variables beyond smoking behaviors. The score is able to identify individuals with the highest risk of disease across smoking statuses.

Cigarette smoking is well-established as the single largest risk factor for COPD. However, a striking proportion of 20%-30% of COPD cases worldwide consists of never smokers^2,3^. Previous studies have investigated the associations between a small set of exposures such as air pollution and occupational exposures (e.g. gas and chemicals)^32–34^ and COPD. However, individuals are simultaneously exposed to a magnitude of multiple factors. Thus, considering broader categories of environmental exposures to assess risk for COPD^28,35^ may be useful to screen populations beyond smoking While smoking status is one of the strongest predictors of COPD, it does not provide granularity for risk stratification. SERS achieved marginally greater predictive ability for COPD in the total population compared to smoking behaviors. However, within smoking status subgroups, SERS was able to stratify low- and high-risk individuals. In never smokers, previous smokers, and current smokers, individuals in the highest quintile of SERS had an HR of 2.33 (95% CI 1.95 to 2.79, P < 0.0001), 4.31 (95% CI 3.62 to 5.14, P < 0.0001) and 5.67 (95% CI 4.62 to 6.95, P < 0.0001), respectively, for developing COPD compared to the remaining population. Furthermore, SERS was able to identify never smokers with a higher risk of COPD than previous or current smokers: never smokers in the highest SERS decile had an HR of 4.95 (95% CI 1.56 to 15.69, P=6.65×10^−3^) and 4.54 (95% CI 2.39 to 8.60, P=4.55×10^−5^) compared to current smokers and previous smokers, respectively, with SERS in the bottom deciles.

We also compared SERS against a composite genome-wide polygenic risk score (PGS) from published weights of 2.5 million SNPS that has been previously demonstrated to be the most predictive genetic risk score for incidence and age of onset of COPD to date^11,13,14^. In our study population, we found that the PGS had significantly lower predictive accuracy than smoking behaviors or SERS in the entire evaluation cohort as well as within each smoking group. PGS was also unable to identify never smokers with an elevated risk of COPD compared to individuals who were previous or current smokers. Second, while SERS and PGS were not significantly correlated with each other, we found that they were additive in the joint model for predicting COPD. We found that having both elevated SERS and PGiS confers a much greater risk for disease compared to having only one elevated score. We also investigated how SERS and PGS may rescue the risk conferred by the other score. Such phenomena are expected under a liability threshold model, in which genetic and environmental effects combine to determine an individual’s total disease liability^36^. Individuals with high SERS and low PGS had an HR of Individuals with high SERS and low PGS had an HR of 4.50 (95% CI 3.08 to 6.57, P < 0.0001) for COPD compared to individuals with low SERS and high PGS, suggesting that the effects of genetic risk on COPD depend on the risk conferred by environmental factors. These results are supported by COPD genetic loci related to nicotinic acetylcholine receptors and smoking-related behaviors (e.g. *CHRNA3* and *AGPHD1*)^37^.

COPD is a disease that has well-documented disparities between groups worldwide. While PGS has been shown to be far more accurate in European than non-European ancestry groups^24^, it was unclear if a similar trend would hold for socioeconomic and environmental factors. Unlike PGS, which decays in accuracy from the study populations as a function of ancestry and genetic distance, SERS performance is driven by cultural, racial, socioenvironmental, and other phenomena. To consistently compare group effects between PGS and SERS, we investigated the generalizability of SERS in predicting COPD risk in non-European ancestry populations by evaluating the performance of SERS in several subsets of non-European ancestry populations in the UK Biobank. The prediction accuracy was consistently lower in the non-European ancestry populations compared to the European evaluation set. In our study population, there were differences in the makeup of some of the most important factors of SERS. For example, CSA, AFR, and EAS ancestry populations had a much smaller proportion of alcohol drinkers and a much higher proportion of being in paid employment compared to the EUR ancestry population. There have also been racial differences in COPD patients^38^. We recognize, however, that the smaller sample size of non-European individuals in the UKB results in lower power and confidence in our conclusions. Furthermore, participants included in the UKB may not be representative of the general population as they tend to be older and are prone to healthy volunteer selection bias^39^. We recognize that an inherent challenge with generalizing results from case-control studies is that ascertainment induces positive correlations between genetic and environmental effects where none may exist in the unascertained population^40^. Our results should be replicated in datasets with more diverse characteristics and ancestry backgrounds, such as the *All of Us* Project^41^.

We note that there are significant limitations with studying exposure data. First, while easy to measure, self-reported exposures may be prone to measurement error and recall bias^42^. In our study, we assumed that these errors occur at random across all variables considered in the SERS. We also excluded exposure variables with >10% missingness, but future studies with more completeness of variables, such as by imputing missing exposure information, would be valuable. Second, one of the most significant challenges in single cohort observational studies such as the UK Biobank is deducing the direction of causality or potential confounding exposure variables. By excluding individuals who at baseline had a past or current diagnosis of COPD, we are more confident that the socioeconomic and environmental risk factors in our study conferred risk for COPD. However, it is possible that some exposures in the SERS (e.g. response to major dietary changes in the past five years) may be explained by other comorbidities. Further studies using causal inference approaches such as Mendelian randomization can better inform the directions of effects between exposures and disease^43–45^. This is particularly relevant when considering scenarios where diagnostic biases are likely, such as whether smokers are more likely to be diagnosed with COPD regardless of underlying genetic liability. Furthermore, understanding the genetic associations is especially important for considering interventions as it can inform biological pathways and mechanisms of COPD. In our study, our model gives the most generous estimate for the PGS as the original GWAS used in developing the PGS contained some overlapping samples from the UK Biobank. Despite this, our estimates of prediction are consistent with previous reports of AUC of between 0.6 and 0.75 for predicting COPD^11^. While the composite PGS we used is based on multi-trait analysis of quantitative spirometry GWAS, it has been demonstrated to be more predictive of COPD cases and time to diagnosis than other existing PGS. It is, however, unclear how much could be improved by considering a multi-trait analysis of genetically correlated traits, such as GWAS of COPD, asthma, and other phenotypes relevant to lung function^46^.

Until recently, studying the cumulative effects of environmental exposures has not been possible on a large scale. With the rise in population-level “biobanks” and high-dimensional epidemiological cohort ‘omics data, there are new opportunities to systematically consider a greater range of non-genetic factors. Leveraging the data available from the UK Biobank, we constructed and validated the first COPD risk score that summarizes the risk conferred by a broad set of socioeconomic factors and non-smoking environmental exposures.

## Data Availability

UK Biobank data are available by application via https://www.ukbiobank.ac.uk/. PXStools is available for download at https://github.com/yixuanh/PXStools. Weights for the composite polygenic risk score were downloaded from https://www.copdconsortium.org/polygenic-risk-score.

https://github.com/yixuanh/PXStools

https://www.copdconsortium.org/polygenic-risk-score

https://www.ukbiobank.ac.uk/

## CONTRIBUTIONS

Y.H. A.R.M, and C.J.P designed the study. Y.H. processed, analyzed, and conducted statistical analysis on the data. Y.H., D.C.Q., J.A.D., M.H.C, and E.K.S interpreted the data. A.R.M. and C.J.P obtained funding. All authors provided critical feedback and revisions for the manuscript.

