## Supplementary Figures for "Prediction and stratification of longitudinal risk for chronic obstructive pulmonary disease across smoking behaviors"

**Supplementary Figure 1:** Manhattan plot of XWAS results. The negative log of the FDR adjusted P value is plotted for each factor. The horizontal line indicates where FDR adjusted P value =0.05. The color represents the exposure category, and exposures are ordered alphabetically.

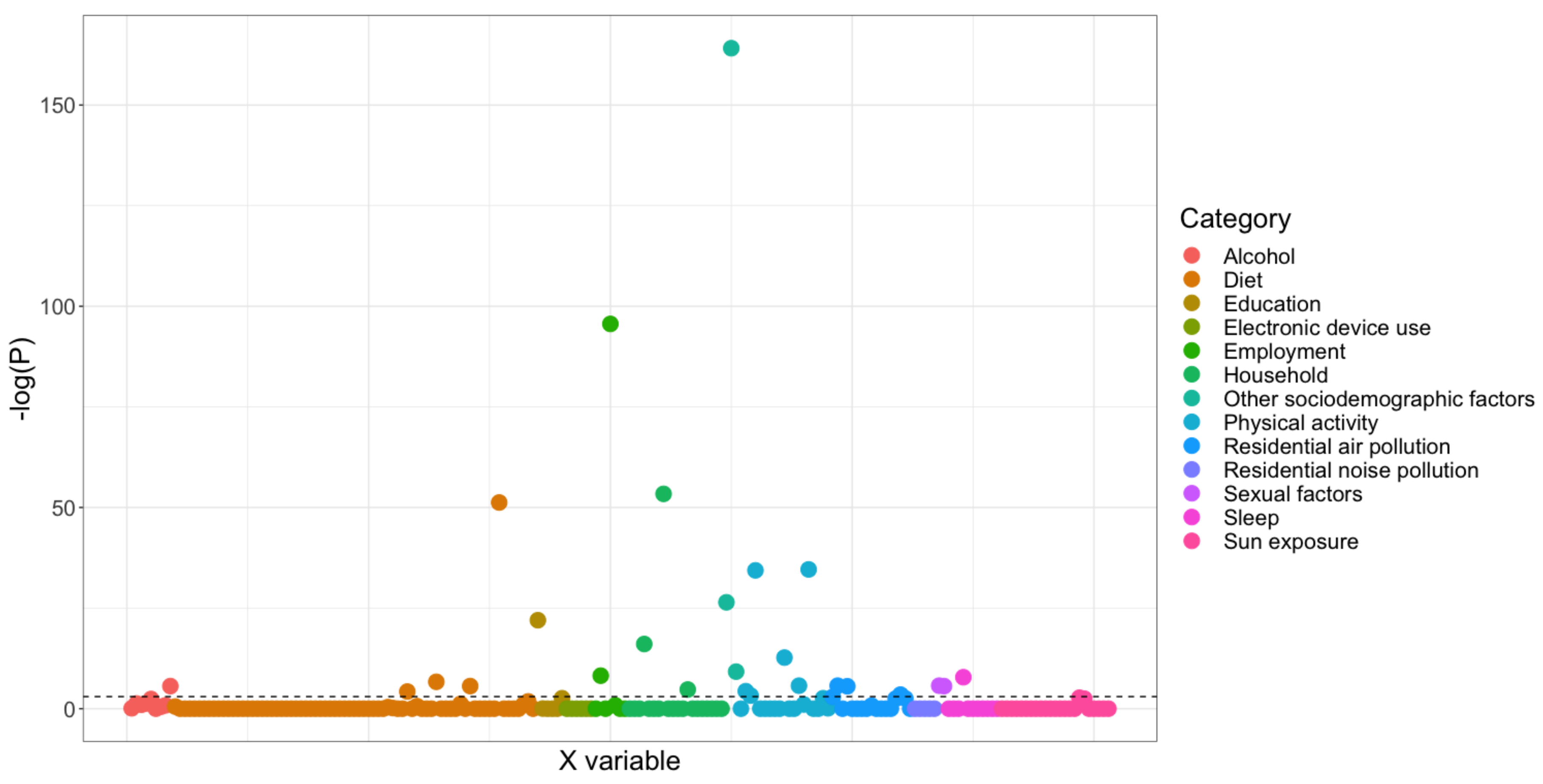

**Supplementary Figure 2**: Cumulative incidence plot stratified by PRS quintile, SERS quintile, and smoking status for EUR evaluation set.

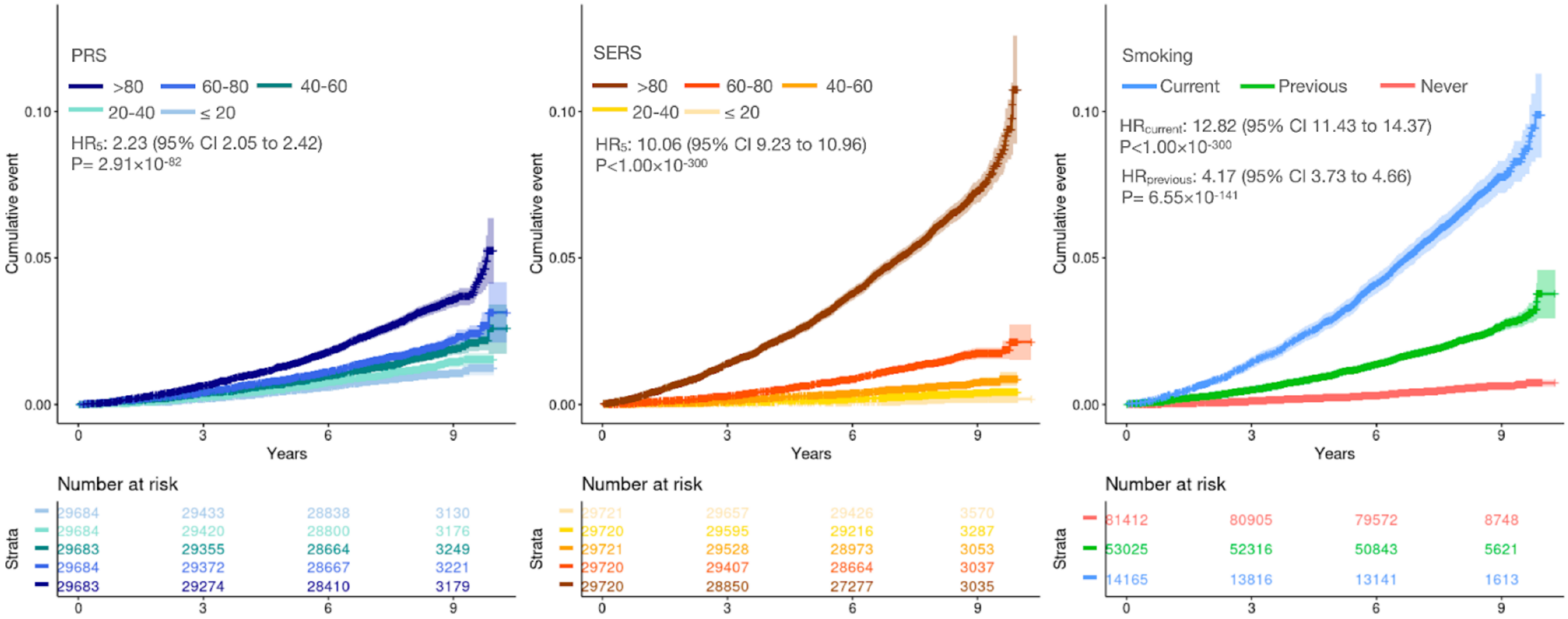

**Supplementary Figure 3:** Stratification of COPD incidence across smoking statuses in the EUR evaluation set. The polygenic (blue) and polyexposure (orange) risk scores were binned into 10 deciles for individuals in each smoking status group. The incidence of COPD for each decile is plotted on the y-axis

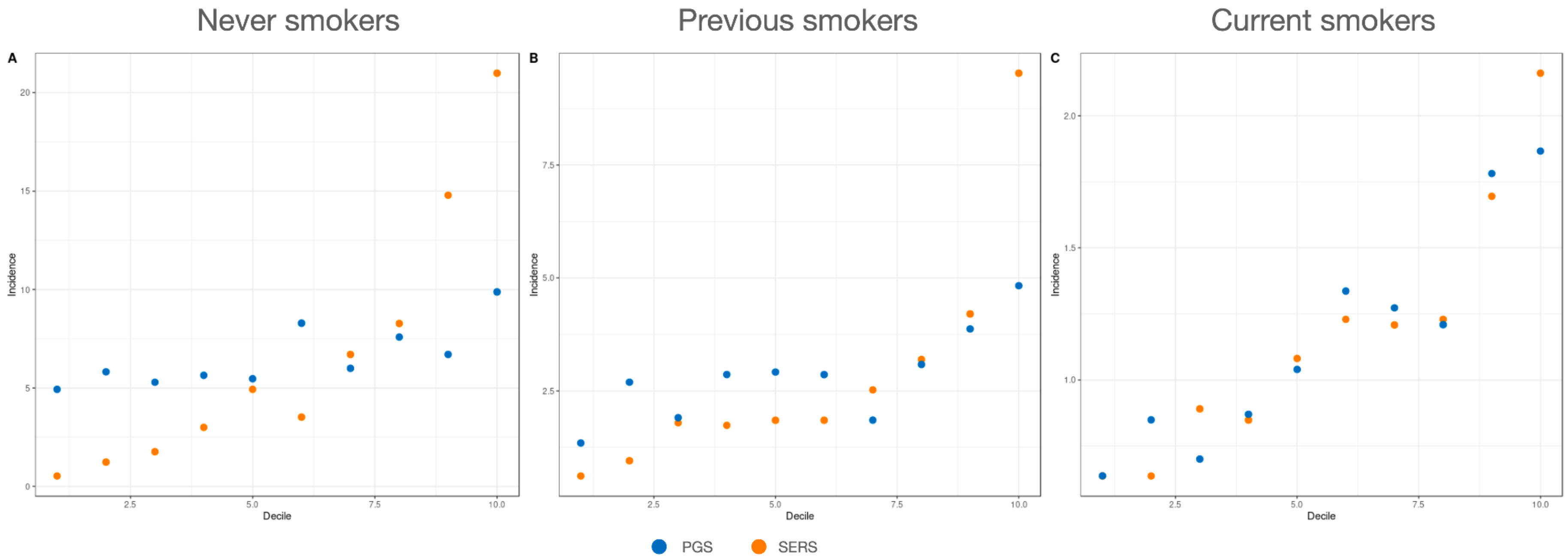

**Supplementary Figure 4:** Distribution of risk scores in EUR individuals with COPD (N=1425). All EUR evaluation set individuals were binned into 100 percentiles based on their SERS or PGS. The distribution of the percentile for individuals who were later diagnosed with COPD is shown.

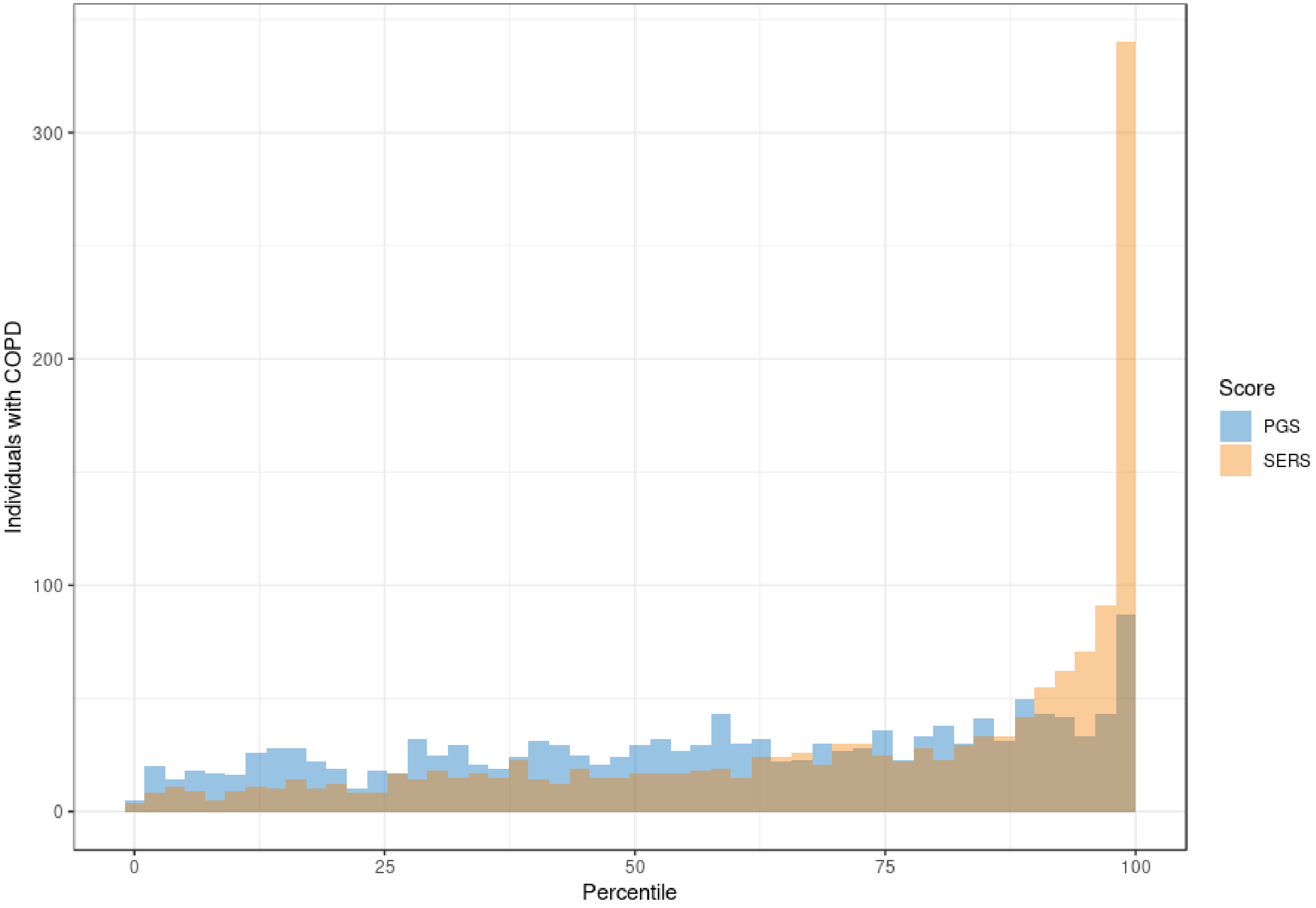

**Supplementary Figure 5:** Average time to event for individuals who are later diagnosed with COPD for each SERS decile.

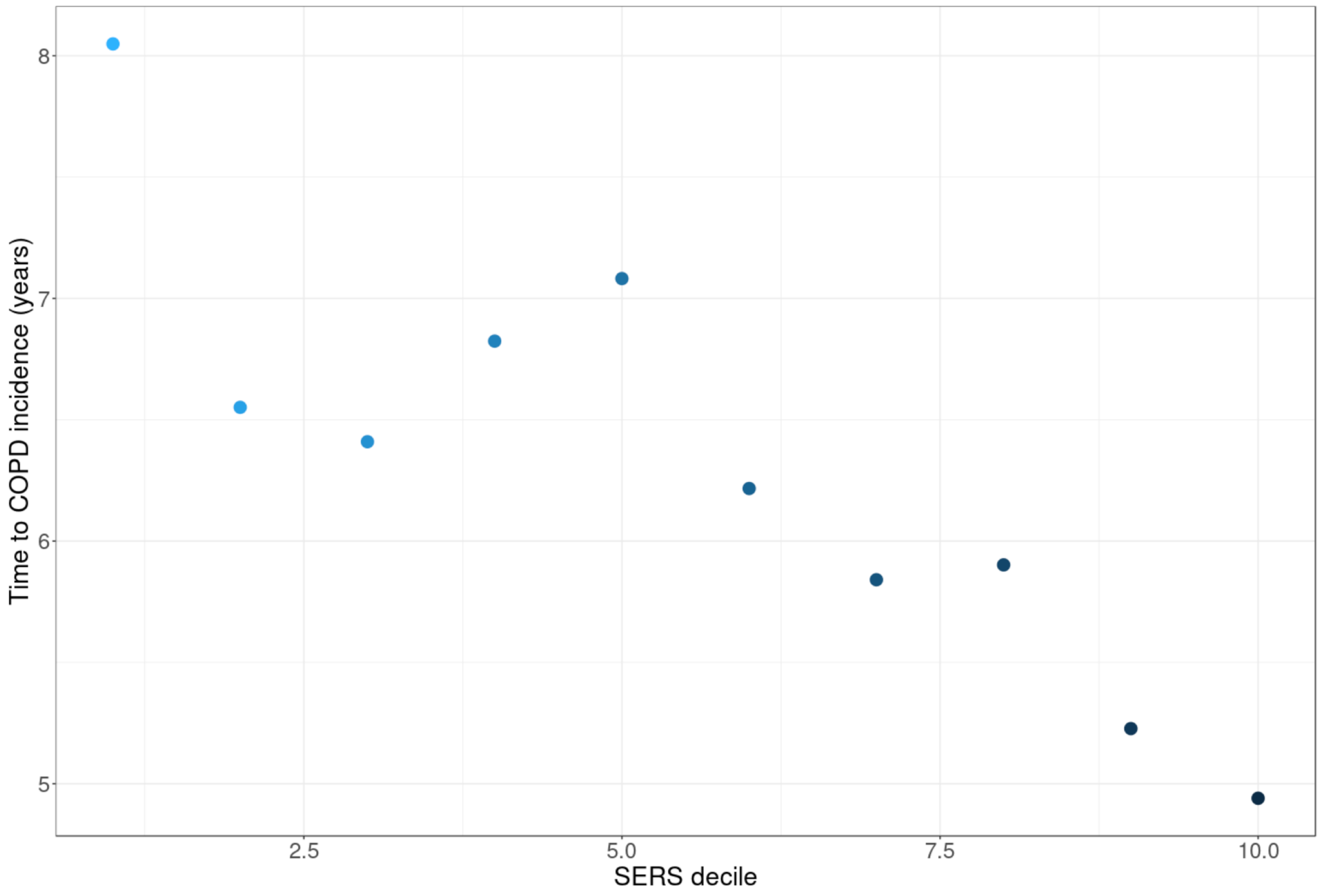

**Supplementary Figure 6:** Cumulative incidence plots for never smokers (left), previous smokers (middle), and current smokers (right) stratified by PGS (blue shades) quintiles. The distribution of PGS for each smoking status is shown in each panel.

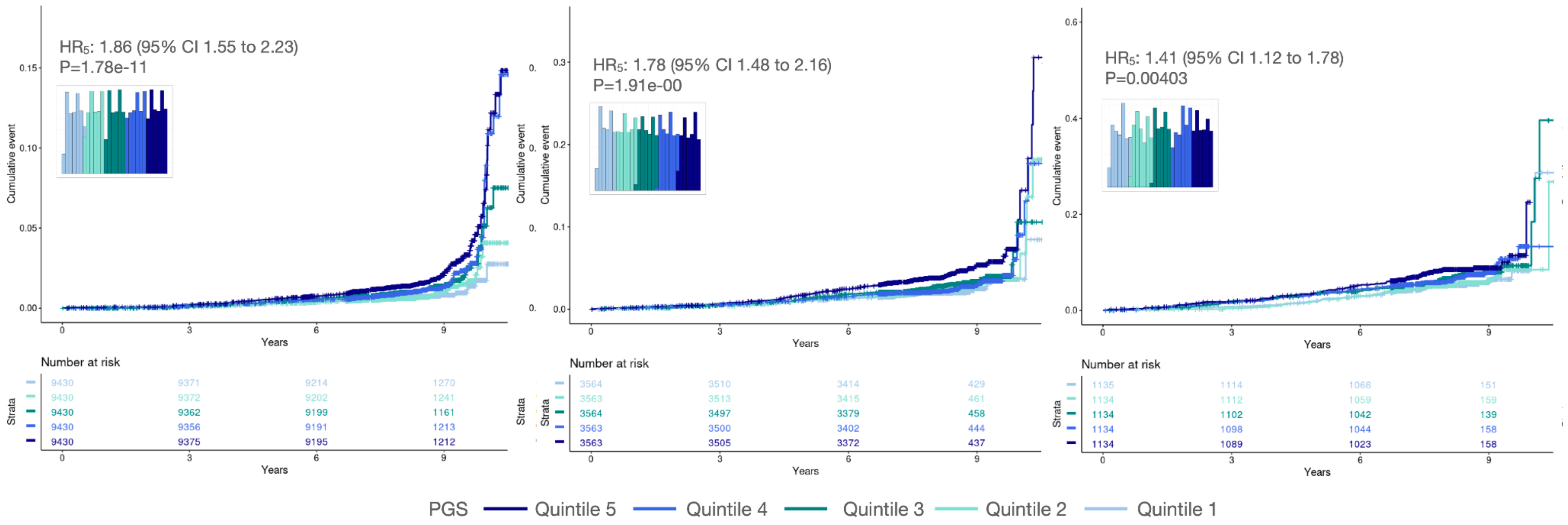

**Supplementary Figure 7:** Cumulative event plot stratified by different combinations of PXS and PGS in the EUR evaluation set. High- and low-risk scores are defined as being in the top and bottom quintiles, respectively. PXSH PGSH: high PXS and high PGS. PXSH PGSL: high PXS and low PGS. PXSL PGSH: low PXS and high PGS. PXSL PGSL: low PXS and low PGS.

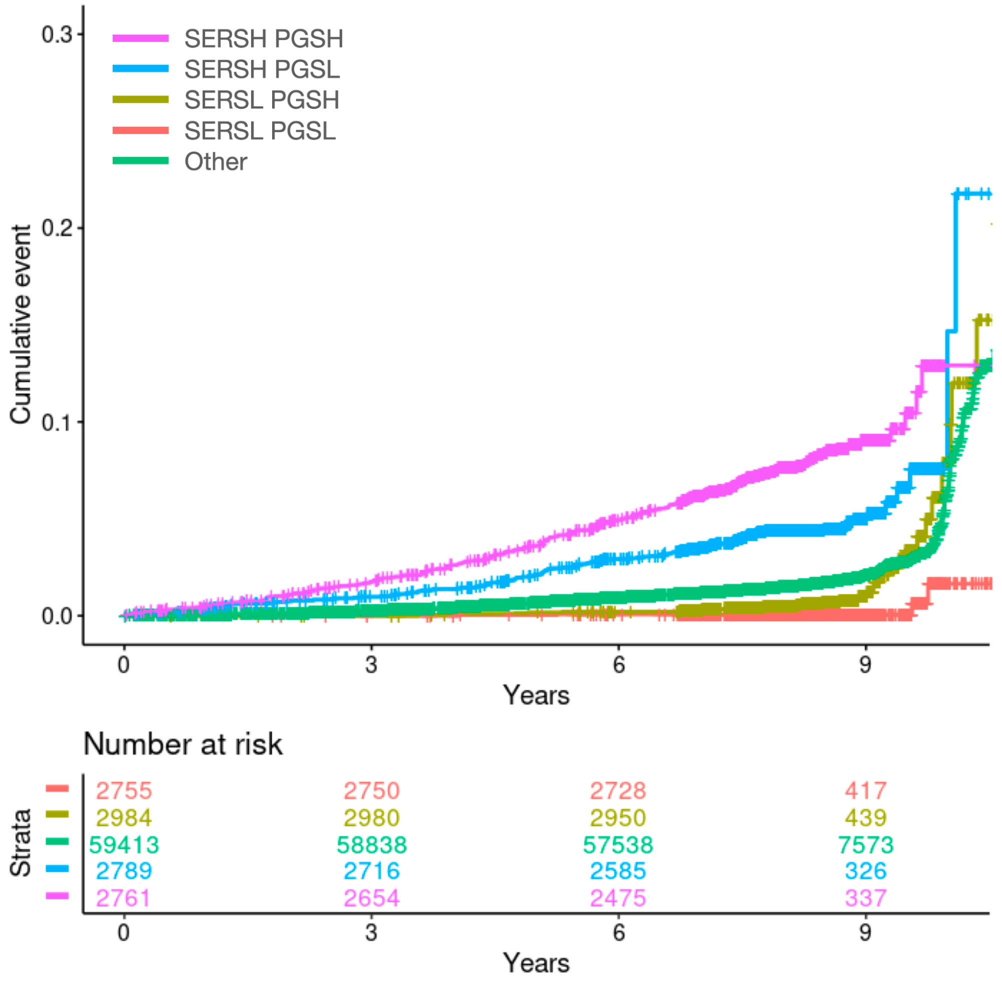

**Supplementary Figure 8**: Venn diagram of individuals with incident COPD who have the highest (top quintile) and lowest (bottom quintile) risk scores in EUR evaluation set. SERSH: highest socioeconomic and economic risk score, PXSL: lowest socioeconomic and economic risk score, PGSH: highest polygenic risk score, PGSL: lowest polygenic risk score.

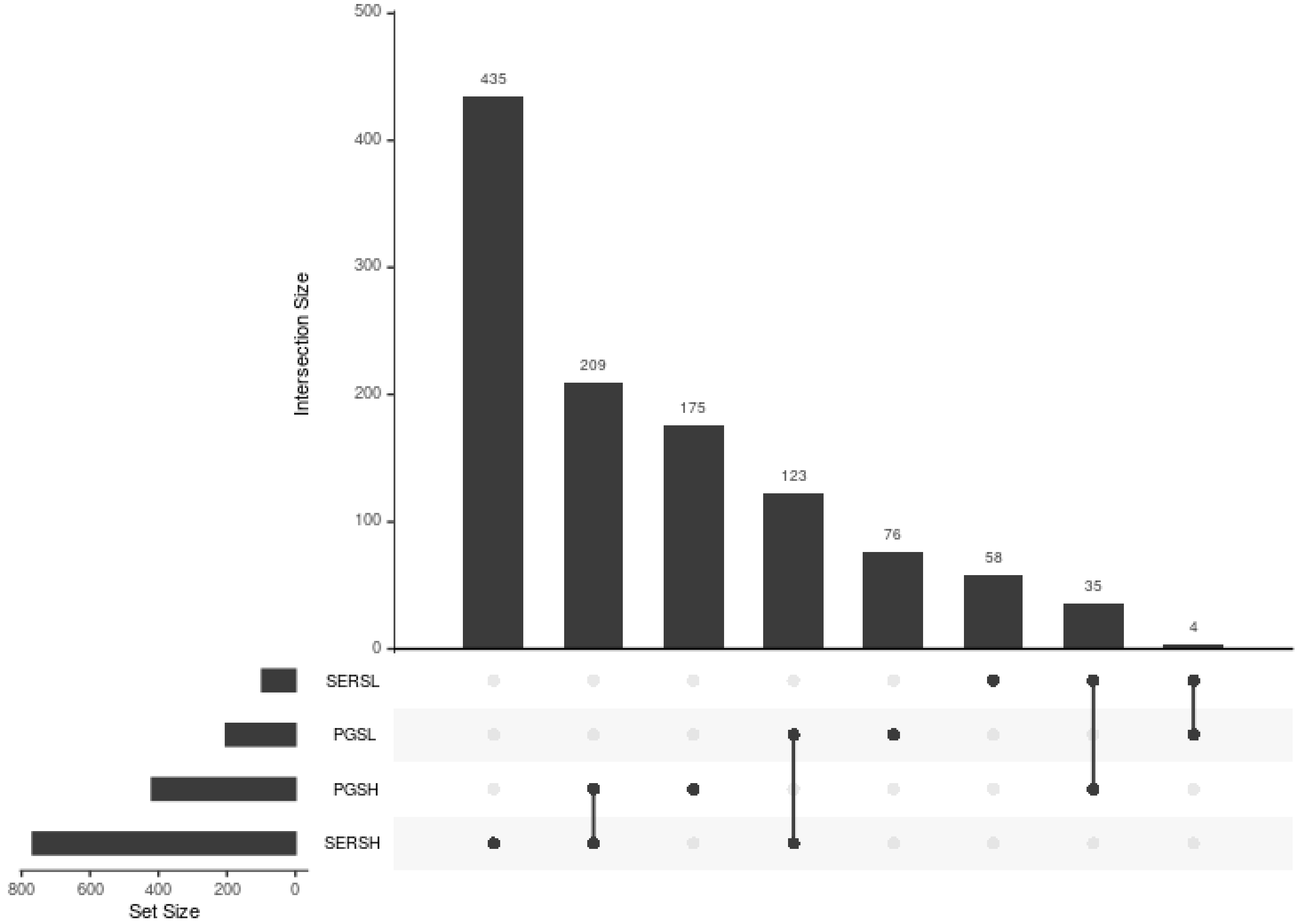
